# An Adaptive, Interacting, Cluster-Based Model Accurately Predicts the Transmission Dynamics of COVID-19

**DOI:** 10.1101/2020.04.21.20074211

**Authors:** R. Ravinder, Sourabh Singh, Suresh Bishnoi, Amreen Jan, Abhinav Sinha, Amit Sharma, Hariprasad Kodamana, N. M. Anoop Krishnan

## Abstract

The SARS-CoV-2 driven disease, COVID-19, is presently a pandemic with increasing human and monetary costs. COVID-19 has put an unexpected and inordinate degree of pressure on healthcare systems of strong and fragile countries alike. In order to launch both containment and mitigation measures, each country requires accurate estimates of COVID-19 incidence as such preparedness allows agencies to plan efficient resource allocation and design control strategies. Here, we have developed a new adaptive, interacting, and cluster-based mathematical model to predict the granular trajectory COVID-19. We have analyzed incidence data from three currently afflicted countries of Italy, the United States of America, and India, and show that our approach predicts state-wise COVID-19 spread for each country with high accuracy. We show that R_0_ as the basic reproduction number exhibits significant spatial and temporal variation in these countries. However, by including a new function for temporal variation of R_0_ in an adaptive fashion, the predictive model provides highly reliable estimates of asymptomatic and undetected COVID-19 patients, both of which are key players in COVID-19 transmission. Our dynamic modeling approach can be applied widely and will provide a new fillip to infectious disease management strategies worldwide.

## Introduction

Since the first reports from China^1–3^, COVID-19 has spread to all the continents resulting in the infection of more than 1.5 million people and a death toll of more than 100,000^4,5^. Due to the severity of the pandemic, many countries have implemented complete or partial lockdowns and international travel restrictions^6–8^ to stem disease transmission^9,10^. As the COVID-19 pandemic presents a very dire economic and humanitarian scenario for most countries worldwide, it is imperative that afflicted governments have ready access to highly reliable estimates of COVID-19 spread across their states and regions. Such predictive incidence data will enable deployment of resource allocation strategies, development of new socio-economic policies and upgradation of healthcare facilities so as to minimize detrimental effects in each country^7,8,11^.

Several studies have modeled the COVID-19 pandemic at the city, state, or country level^6,8,12–14^ using the common Susceptible–Exposed–Infected–Removed (SEIR) model^15^ that can capture the dynamics of an infectious disease such as COVID-19. In this model, the population is divided into four categories of which “susceptible” individuals may become “exposed” to the virus through “infected” people who will eventually be “removed” (that is, they can no longer infect others). Removed population refers to the individuals who have recovered or died. The traditional SEIR model when applied to model COVID-19, however, suffers from the following two major limitations: (i) it assumes homogeneity in a large population via keeping the basic reproduction number R_0_ a constant (i.e., local variations in the transmission dynamics within a large population are not accounted for) ^15–17^, and (ii) it assumes a “closed population” without demographic variation stemming from births, deaths or migration^15^.

China reported its first case on 31 December 2019, with a peak in cumulative cases in an eight-week interval and thence a plateauing. Italy followed the same trajectory after ∼11 weeks and then the USA after ∼13 weeks (of the first case in China). In India, cases rose after ∼12 weeks of the first case in China, and although both cases and deaths are still on the rise in the USA and India, Italy is already witnessing a decrease in daily new cases. To understand the trends of this epidemic, many studies in different countries have employed the R_0_ that was estimated from China. As in other directly contagious diseases, COVID-19 spreads primarily due to human transmission of the pathogen (coronavirus) from city-to-city, or state-to-state, or country-to-country, and this involves significant migration of humans^6,12,13^. The dynamics of disease spread, therefore, involves a few primary cases and an index case up to which point the R_0_ is limited in its value. Beyond this, when the infection starts to move from index cases to their contacts, the R_0_ assumes greater magnitude and then it can drive community transmission that is currently being witnessed in many countries and feared in others that are behind in their epidemic evolution.

Although R_0_ is a measure of communicability of COVID-19, its upper range determines the speed of spread. Estimation of R_0_ assumes that everyone around a primary case is equally susceptible to the infection and thereby suggests that it is dependent on the causative agent alone. However, R_0_ is a function of direct and indirect interactions between the agent, host and environment. The hosts’ immune status, genetic makeup, comorbidities, gender and smoking can contribute towards disease transmission. Equally, the environment that supports transmission is dynamic via variations in temperature, humidity, population density, migration, adaptive interventions like quarantine/isolation/social distancing, socio-economic conditions and so on^18–22^. Hence, the use of a constant value for R_0_ cannot capture the evolving transmission dynamics accurately. To address this challenge, we first estimated the spatio-temporal variations of R_0_ in Italy, USA and India (see Figure 1). Specifically, we tracked COVID-19 spread in each state/region within these countries and then computed R_0_ by explicitly solving the SEIR equations. Interestingly, we observed that R_0_ exhibited significant spatial and temporal variations (see Figure 1), and hence it was deemed inappropriate to be used as a constant for any large population.

**Figure 1.**
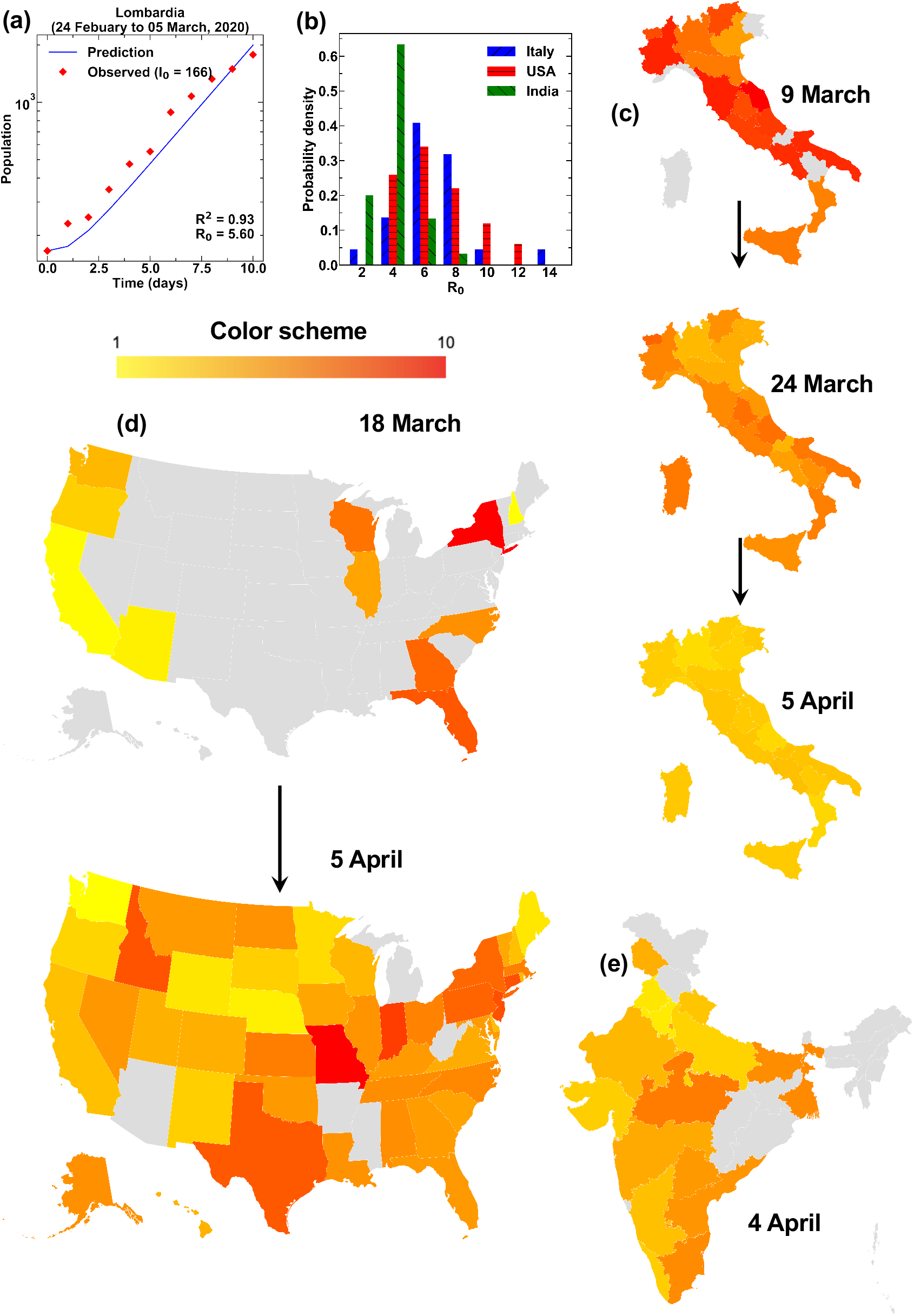
Basic reproduction number R_0_. **(a)** SEIR model fitted against the observed data (from 24 February 2020 to 9 March 2020) for Lombardia (Italy) to compute its R_0_. Similar approach was applied to all the states for different time periods (see Supplementary Material). **(b)** Histogram of R_0_ values for Italy (24 February to 9 March), USA (4 March to 18 March), and India (10 March to 24 March) in the early stages of the COVID-19 pandemic. **(c)** R_0_ in different regions of Italy on 9 March, 24 March and 5 April 2020. **(d)** R_0_ in different states of the USA on 18 March and 5 April 2020. **(e)** R_0_ in different states of India on 4 April 2020. The coloring scheme for **(c), (d)**, and **(e)** is common and is shown in the legend. Grey regions represent the states for which R_0_ cannot be estimated reliably due to the low number of cases.

To address the granularity in R_0_, we developed a new adaptive, interacting, cluster-based SEIR (AICSEIR) model that, we show, can capture the transmission dynamics of the COVID-19 pandemic within a heterogeneous population to high accuracy (Figure 2). Hereon, the term state represents a subpopulation (or a cluster) in a country. State, therefore, corresponds to the geo-administrative boundaries within India and the USA, and regions in Italy. Our new model divided any given country’s entire population into multiple, interacting clusters that mingled stochastically. This enabled us to predict the trajectories of COVID-19 transmission in three heterogeneous populations of Italy, USA, and India up to the state/region level. Typically, R_0_ is estimated by fitting an exponential curve in the early infection stages following the assumption that 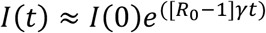. However, due to the paucity of new cases in the early phases, the dynamics can be highly stochastic and influenced by large, noisy fluctuations, which together cause R_0_ estimates to be unreliable^15,16,23^. By the time stochastic fluctuations become negligible, the epidemic behavior will tend to be nonlinear due to recoveries or deaths in infected populations rendering the exponential approximation invalid^15^. In such cases, the exponential approach will lead to a significant underestimation of R_0_ due to the removed population (as it is not accounted for in the exponential model). To address these caveats, we computed R_0_ by optimizing predictions from the SEIR model for each state within a country as a function of time (see Methods). This new approach is able to capture the time dynamics of R_0_ that emanate as a result of public health interventions in a given country. In particular, we decided to re-estimate R_0_ on a fortnightly interval in order to capture its variability.

**Figure 2.**
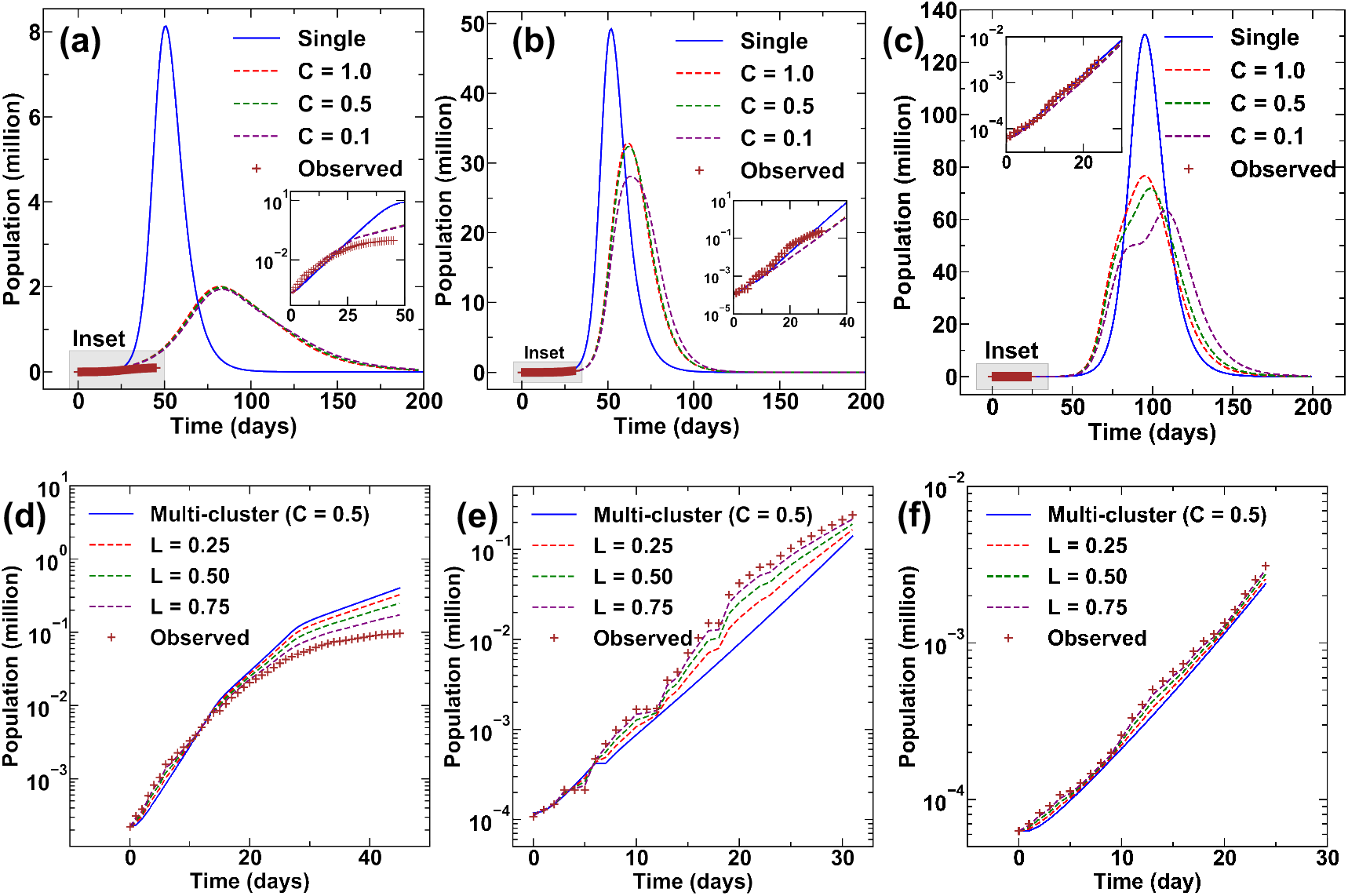
Countrywide spread of COVID-19. Evolution of the pandemic in (a) Italy (b) the USA and (c) India with respect to time. This is based on the traditional SEIR (single cluster) and AICSEIR models with *C* = 1.0, 0.5, 0.1. *C* represents the inter-cluster mobility of the population where *C* = 0 represents zero mobility and *C* = 1 representing restriction-free mobility. INSET for (a), (b), and (c) show fit of model predictions and observed infected cases (square markers). We noted that the variance in comparison to the mean trajectory is significantly small, and it was hence omitted in these figures. The best estimates considering the error between model and observation for (c) Italy, (d) the USA, and (e) India with *L* = 0.25, 0.50, and 0.75. Note that a lower value of *L* suggests increased confidence in the observation, while a higher value of *L* suggests increased confidence in the model. Time *T* = 0 corresponds to 24 February 2020 for Italy, 4 March 2020 for the USA and 10 March 2020 for India.

## Methodology

### (i) Dataset

The datasets used for the study include the following. (i) The total number of COVID-19 active, and removed cases in three countries—Italy, the USA, and India, along with the state-/region-wise details. These data are obtained from the WHO and the respective government databases^4,24–29^. (ii) Population data of each of the states-/regions in the three countries. (iii) Distance between the capital cities of the states in each of the countries is directly calculated from the latitude and longitude of the respective cities. Complete data used in the study are provided in the Supplementary Material.

### (ii) Adaptive interacting cluster-based SEIR (AICSEIR) model

Herein, we present the proposed AICSEIR model (Eq. (1) – Eq. (8)), developed by suitably extending the heterogeneous SIR model^15^ that captures the coupling dynamics between populations residing at different geographical locations:

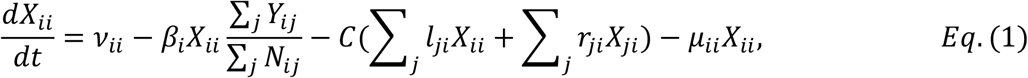

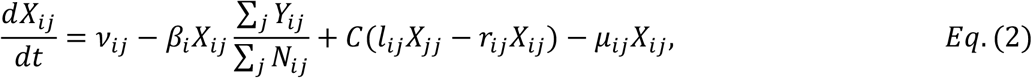

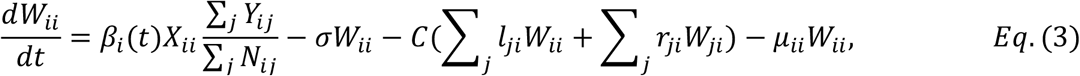

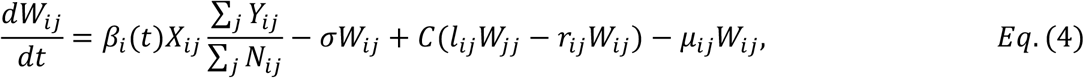

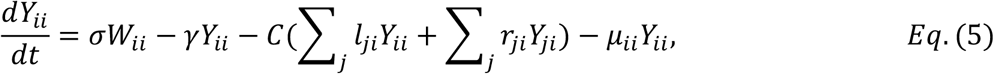

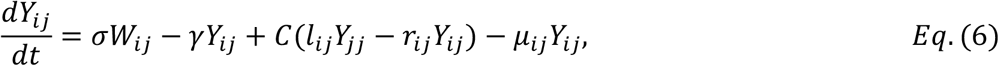

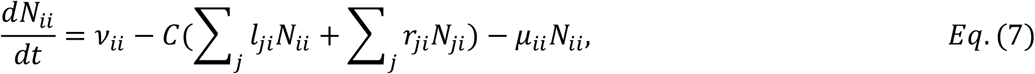

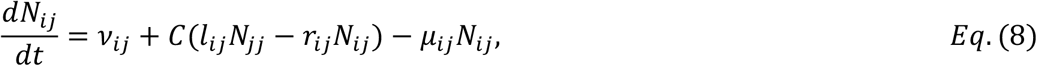

In the above equations, *X*_*ii*_, *Y*_*ii*_, *W*_*ii*_, *N*_*ii*_, *ν*_*ii*_, *μ*_*ii*_ denote the number of susceptible, infected, exposed, total hosts, births, and deaths, respectively, in a subpopulation (cluster) ‘*i*’ that live in subpopulation ‘*i*’ and *X*_*ij*_, *Y*_*ij*_, *W*_*ij*_, *N*_*ij*_, *ν*_*ij*_, *μ*_*ij*_ denote the number of susceptible, infected, exposed, total hosts, births, and deaths in subpopulation ‘*i*’ that live in subpopulation ‘*j*’, respectively. In this study, it is assumed that the number of births and deaths compared to the number of susceptible, infected, exposed, total hosts are negligibly small for the time-period considered and therefore set to zero.

The parameter *γ* is called the removal or recovery rate, defined as the reciprocal of the average infectious period. In this study, the average infectious period is considered to be three days. *β*_*i*_(*t*) the parameter indicates the cluster-wise spread of the disease as a function of time. *β*_*i*_ (*t*)is evaluated as *β*_*i*_(*t*) = *γR*_*i*0_(*t*) where *R*_*i*0_(*t*) is the time-varying basic reproductive ratio, a key measure that governs the spread of the epidemic. *σ* parameter is the inverse of the average latent period or average incubation period. In this study, the average incubation period is assumed to be seven days^8,30^.

The variable *l*_*ij*_ measures the rate at which individuals leave their home population ‘*j*’ and to subpopulation ‘*i*’, and *r*_*ij*_ measures the rate at which individuals leave the subpopulation *‘i’* and to their home population ‘*j*’. We have assumed that during the onset of an epidemic, any individual in the home population would choose to stay there and a fraction of the individuals that live in population ‘*i*’, may return to their home population *‘j’*. Therefore, we have considered *l*_*ij*_ to be zero in the model, while *r*_*ij*_ is modeled as a stochastic parameter. To this extent, we have assumed that the fraction of the home going migrant population from each subpopulation ‘*j*’ per day will be capped to a fraction ‘*frac*’ of the subpopulation. Hence, the matrix *r* is generated as a *S* × *S* matrix, where *S* denotes the total number of states in a country, with each element *r*_*ij*_ is sampled from *r*_*ij*_ ∼ *U*[0, *frac*], where *U* is the Uniform distribution, with a restriction of *max*(*r*_*ij*_) = *frac*. In the study, without loss of generality, *frac* is set to be 0.10.

Once *r*_*ij*_ is frozen, the next step is to calculate *X*_*ii*_ and *X*_*ij*_. This involves the allocation of the home going migrant population from a native subpopulation to (*s* − 1) other native subpopulations. To this extent, we have assumed that the home of the migrant population is distributed to (*s* − 1) other subpopulations in a ratio directly proportional to the population of the receiver state and inversely proportional to the distance between them. Further, for simplicity, we assume the state capitals are the point of entry and exit points of the migrant population. If we denote *S*_*i*_ be the total population of state *i*, then *X*_*ii*_ = (1 − *r*_*ii*_)*S*_*i*_ and 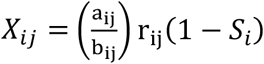, where a_ij_ is the fraction of the population of the receiver state normalized with the population of remaining (*s* − 1) states and b_ij_ is the fraction distance between capital cities from the feeder state’s capital normalized with distance to the capital cities of the remaining (*s* − 1) states.

The infected population matrix *Y* is initialized with *Y*_*ii*_ is equal to the actual number of cases reported in the state *i* at the start of the simulation day and *Y*_*ij*_ set to zero for all the states. Also, the exposed population matrix *W* is initialized identically to that of the infected population matrix *Y* to start the simulation. Further, we add an inter-cluster restriction parameter *C* to tune effect of restrictions imposed, as the result of various interventions enforced by the state/central administrations, on the mobility of the migrant population from feeder state to receiver state with *C* = 0 representing zero mobility, and *C* = 1 representing restriction-free mobility.

### (iii) Computation of R_0_

In this study, *R*_0_ is computed by directly fitting the observations to the proposed model by minimizing the prediction of infections. The optimization formulation for computing *R*_0_is given below:

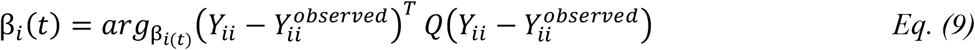

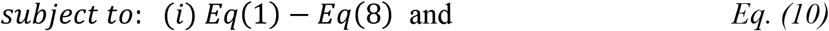

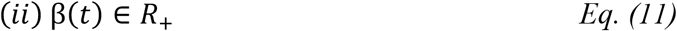

Here, 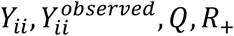 are infections predicted by the model, observed infections, a suitable weight, and a set of real numbers, respectively. Once β_*i*_(*t*) is computed, *R*_*i*0_(*t*) *is obtained as β*_*i*_(*t*) = *γR*_*i*0_(*t*). However, a key point is that due to various interventions of state-wise and country-wise interventions *R*_*i*0_(*t*) would be varying over time. Hence, to make our study realistic, we adaptively re-estimate *R*_*i*0_(*t*) using every 14 days data by employing Eq. (9)–Eq. (11).

### (iv) Model correction using real-time observations

It is imperative to reconcile the model predictions of AICSEIR model with clinically diagnosed infected case due to the following reasons: (i) Model predictions will be overestimating the total number of infected cases as predictions only depend on *R*_0_ and the initial infected population. (ii) Clinically diagnosed cases will be underestimating the total number of infected cases due to the testing limits or saturation. Hence, a realistic estimate of the total number of infected cases will be following a middle ground between the two. To this extent, we propose a weighted prediction correction strategy motivated by Kalman filter estimates:

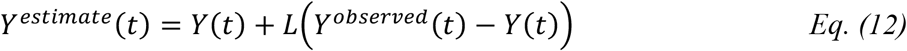

Here, *Y*^*observed*^(*t*) is the clinically diagnosed infected cases, *Y*^*estimate*^(*t*) is a realistic estimate of infected cases, and *L* is the weighting factor with |*L*| ∈ [0,1] and can be tuned based on the real scenarios. *L* value of 0 implies 100% confidence in the model, while an *L* value of 1 implies 100% confidence in the observation^31^.

## Results

### (i) Basic reproduction number of COVID-19

To validate our approach, we used the SEIR model to fit actual COVID-19 incidence data for Lombardia of Italy (Figure 1(a), see Methods), and then computed its R_0_ values^4,24–29^. The high R^2^ value associated with the fit suggests that the derived R_0_ values are reliable for the time-period considered (Figure 1(a) and Supplementary Material). We then proceeded to do this for all the 30 states within India, 45 within the USA and 20 regions of Italy (Figures 1(b)–(e)). While in few cases, the R^2^ fits were poor due to low initial infection load, most states in the three countries produced highly reliable R_0_ values (Figures 1(c)–(e) and Supplementary Material). It was noted that states with high incidence returned very robust R^2^ values and thus, we considered all R_0_ values with R^2^ > 0.8. For the few other states, R_0_ was assumed to be the country average. Such analyses resulted in a dynamic R_0_ profile for each of the three countries in the early stages of the COVID-19 outbreak (Figure 1(b)). Interestingly, we observed that for both Italy and the USA, the R_0_ values exhibited significantly broader distribution ranging from ∼2-14 and ∼4-12, respectively (detailed state-wise plots for estimating R_0_ along with the exact R_0_ scores are provided as Supplementary Material). On the contrary, in the case of India, we observed that R_0_ values ranged from ∼ 2-6 (Figure 1(b)). This evident variation in the ranges of R_0_ values is in congruence with the observed slower rate of early COVID-19 spread in India when compared to the USA and Italy despite the fact that all three countries reported their first COVID-19 case at the end of January 2020.

We next analyzed the temporal variations in R_0_ as it is significantly altered due to many factors, including travel restrictions, state-wise lockdowns (as in part of USA) and countrywide lockdown (as for Italy and India). We, therefore, calculated R_0_ for Italy prior to lockdown (that is before 9 March 2020), two weeks into lockdown and four weeks into lockdown (Figure 1(c)). For the USA, we estimated R_0_ with a two-week interval period (Figure 1(d)). Moreover, in the case of India, due to the delayed onset of the spread of disease, we computed a single R_0_ (Figure 1(e)). These data provide the R_0_ landscape as a choropleth map for each country (Figure 1(c)–(e). As is evident, the R_0_ for Italy decreased significantly due to its lockdown routines (Figure 1(c)). Indeed, enforcement of stricter mobility restrictions has reduced Italian R_0_ values closer to unity thereby controlling the growth of the epidemic (Figure 1(c)). For the USA, it is clear that only the states that implemented substantial restrictions have managed to reduce their R_0_ values (Figure 1(d)). For India, the strict screening of incoming international travelers and early imposition of lockdown resulted in reduced R_0_ values in comparison to Italy and the USA. These analyses therefore immediately reveal the benefits of public health interventions, and such modeling approaches may be used widely and routinely for assessment of intervention outcomes.

### (ii) Adaptive interacting cluster-based SEIR (AICSEIR) model

Based on revised R_0_ profiles, we then used our AICSEIR model (see Methods for details) to predict COVID-19 spread in Italy, the USA, and India. For this, our model required total state population, values of distance between the capital cities of two-states, initial infected number (it could be zero) and the temporal variations in R_0_ (as estimated in the previous section, see Methods). The total population of any state was divided into native and migrant categories (latter was set to 10%). It was assumed that the distribution of a state’s migrants was directly proportional to the population of the home state and was inversely proportional to the inter-capital distance. Therefore, two implicit assumptions in these analyses are: (a) people are prone to migration from a highly populated state, and (b) the likelihood of choosing a nearby state for migration is higher. Further, indirect measures of migration, such as airline/train/bus data and the number of tourists, were ignored.

We then compared directly the trajectories of infection prevalence in Italy, the USA, and India using both the traditional SEIR model (represented as single in Figure. 2(a)–(c)) and our new AICSEIR model (Figure 2(a)–(c)). A new parameter C was introduced wherein values of 1.0, 0.5, and 0.1 represent the inter-cluster interaction restrictions (C of 0 and 1 denote the absence of migration versus free migration, see Methods for details). All presented models were run extensively with multiple random seed values to account for the stochastic parameter *r*_*ij*_ that models migration as a random event (see Methods). Using this, a direct comparison of the predictive robustness of SEIR and AICSEIR models in the context of true incidence in the three countries is possible (Figure 2(a)–(c)). We observed SEIR significantly overestimates the peak-infected population (five-fold for Italy and up to 1.8 fold the USA and India). In contrast, the AICSEIR provided a significantly closer estimation of infected cases (Figure 2(a)–(c)). Thus, our approach was able to recapitulate the epidemiological trends both qualitatively and quantitatively not only on a countrywide scale but also for its constituent states/regions.

It is noteworthy that the model provides a prediction for total infected, but the observations are based on clinically detected cases. Therefore, both these estimates suffer from the following deficiencies. The clinically detected cases will always underestimate the number of infected cases as the number of tests conducted limits the detection. Besides, all asymptomatic infections shall be missed. On the other hand, our model might still overestimate the total number of cases (but not as much as the SEIR approach) as it is based on the initial conditions and infection dynamics as per R_0_ values. Indeed, there are a host of other confounding factors that can govern R_0_ such as the climatic conditions, host genetics, immune status, age, gender and co-morbidities. Therefore, the best estimate of total infected population lies between model predictions and actual observation (Figure (d)–(f)). While their difference could be small in the early stages, the disparity could be staggering at later stages. To account for this unreliability, we have added a model correction factor *L*, inspired by the Kalman filter that provides an estimate of the infected population^33^. Here, the estimate of the infected population at any time *t* is computed as the sum of the infected population in the previous timestep *t* − 1 and the difference between observed and model prediction at *t* weighted with *L* (see Methods). |*L*| resides between 0 and 1 based on the confidence of the model and observation: *L* value of 0 implies 100% confidence in the model, while a value of 1 implies 100% confidence in the observation. We suggest that former (*L* = 0) can be used in countries with a scarce level of COVID-19 testing while the latter (*L =* 1) can be used where there is ample testing capacity (Figures 2(d)–(f)). In this scenario, the real observations provide a lower bound of the infected cases while our AICSEIR model provides the upper bound. This, in turn, allows the estimation of infections that may be undetected or asymptomatic, as both play major roles in the transmission of the infections.

### (iii) Representative state-wise prediction of COVID-19

Another facet of our AICSEIR model is its ability to predict the evolution of the infection in state-wise or in clusters. Indeed, the country-wise predictions were computed as the summation of sub-populations (state-wise). To validate further, we selected two states from each country and mapped their COVID-19 burden (Figure 3). The initial, exposed, infected, and removed populations of Calabria and Veneto (Italy), Idaho and Washington (USA), Madhya Pradesh and Uttar Pradesh (India) were assessed (Figure 3). Note that for each country, at least one state chosen had zero initial infected population. For the initiation of infection in these virgin territories, the importation of infected persons would be required based on the cluster interaction term *C* (*C* = 0 would maintain zero infection). We observed that infection trajectories predicted by the model were in excellent agreement with the observed cases for states with zero initial infected population and finite infected population. In other words, through the cluster interaction term, the model is able to realistically predict the spread of COVID-19. We have provided detailed state-wise mapping of populations likely to be infected in future for each state in each of the countries (30 in India, 45 in the USA, and 20 Italy, Supplementary Material). These data will facilitate state-level and national authorities to devise plans for the allocation of public health resources judiciously at a granularity that addresses state-wise disease burden.

**Figure 3.**
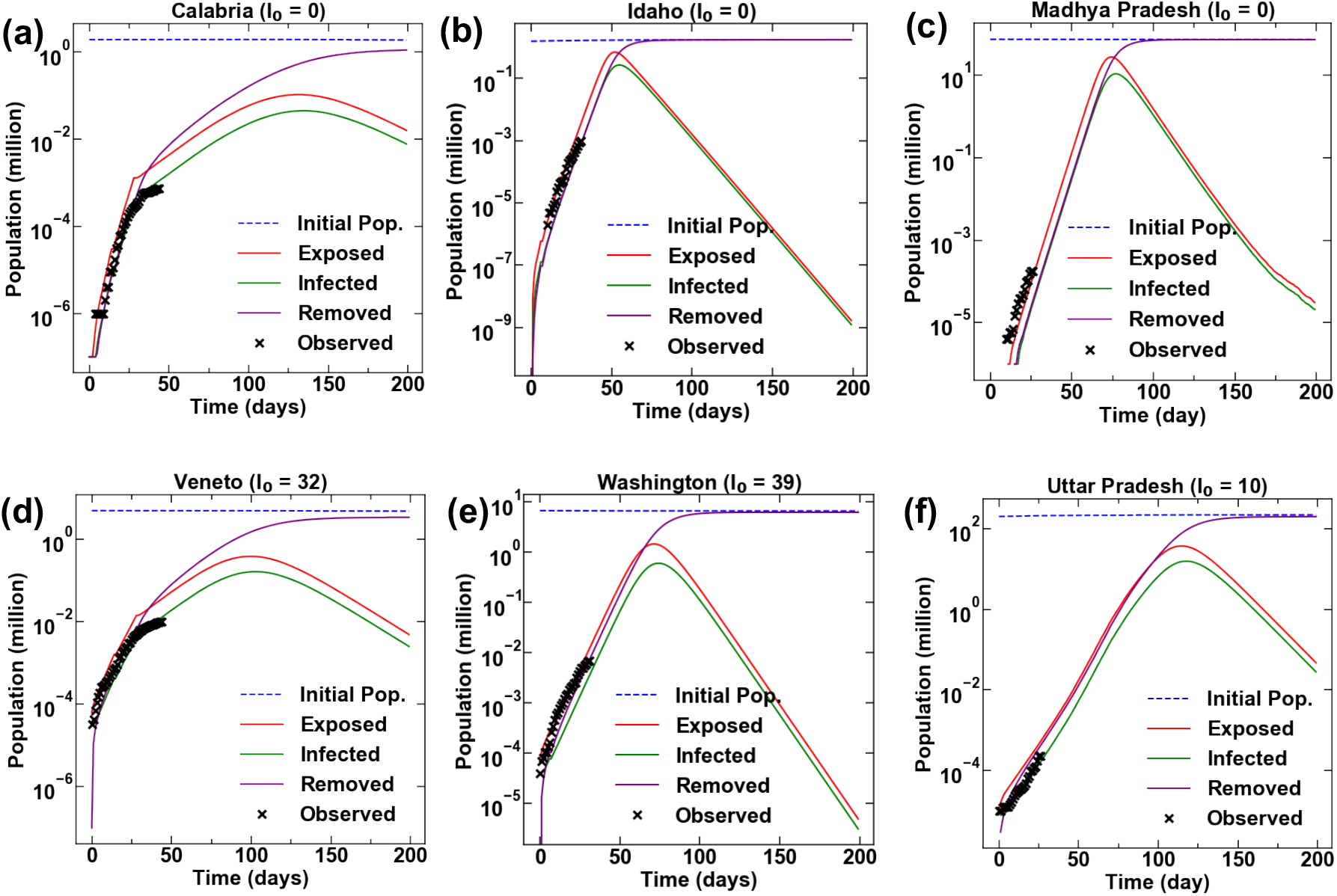
State-wise evolution of COVID-19. Mapping of the pandemic in three states (a) Calabria (Italy), (b) Idaho (the USA), and (c) Madhya Pradesh (India) with zero initial infections as predicted by AICSEIR model in comparison to the observed data. Progression of COVID-19 in three states (d) Veneto (Italy), (e) Washington (the USA), and (f) Uttar Pradesh (India) with non-zero initial infections. It is noteworthy that in both scenarios, our model is able to predict the observed trends to high statistical reliability.

## Conclusion

To our knowledge, previous studies on the COVID-19 pandemic have used a constant value of R_0_ to assess disease spread ^6,8,12,32^. We have clearly demonstrated that R_0_ is not constant and indeed exhibits significant spatio-temporal variations. These fluctuations in R_0_ need to be incorporated in the development of robust and realistic epidemiological models. We show the utility of the SEIR model for estimating R_0_ wherein a simple exponential fit may, in the best case, lead to over-/under estimation of R_0_, and in the worst case, may simply be not valid due to the non-linear variations in disease spread. We propose that temporal variations in R_0_ should be included in an adaptive fashion, while the spatial variations should be included in a granular, cluster-wise model. This approach is capable of capturing realistic infection dynamics across each nation or indeed worldwide. AICSEIR with its tunable interaction parameters, can indeed be applied to other infectious diseases.

There are several outcomes of immediate public health value from our work: (i) we provide robust estimates of infection burden with timelines and this will facilitate proactive development of resource allocation strategies locally^33,34^, (ii) our model provides a caution for regions with low caseload presently as they are likely to follow trends of other highly affected areas in the absence of substantial mobility restrictions, (iii) we suggest a locally graded contextual interventional responses that can factor socio-economic factors and morbidity (note that complete longer-term lockdowns will have notable detrimental economic fallouts resulting in exaggerated impacts on society), (iv) our revised novel coronavirus burden estimates will help map the true extent of infection that includes undetected cases and asymptomatic infections. Although epidemic prediction models tend to discount pivotal contributions from the host and environmental confounders^35,36^, two useful extrapolations of our model are to assess case volumes that may require intensive care and to calculate the true case fatality rates (CFR)^37,38^. The AICSEIR model can thus serve as a valuable tool for strategizing containment and for stemming mortality associated with the COVID-19 pandemic.

## Data Availability

All the data associated with the work is available from the corresponding author upon reasonable request.

## Acknowledgements

The authors thank IIT Delhi HPC facility for computational resources.

## Code Availability

All the codes used in the present work are developed in-house in the python environment and are made available in the GitHub repository: https://github.com/m3rg-repo/COVID_modeling.

## References

1. Wu, F. et al.. A new coronavirus associated with human respiratory disease in China. Nature 579, 265–269 (2020).

2. Zhou, P. et al.. A pneumonia outbreak associated with a new coronavirus of probable bat origin. Nature 579, 270–273 (2020).

3. Zhu, N. et al.. A Novel Coronavirus from Patients with Pneumonia in China, 2019. New England Journal of Medicine 382, 727–733 (2020).

4. Novel Coronavirus (2019-nCoV) situation reports. https://www.who.int/emergencies/diseases/novel-coronavirus-2019/situation-reports.

5. Wu, Z. & McGoogan, J. M. Characteristics of and Important Lessons From the Coronavirus Disease 2019 (COVID-19) Outbreak in China: Summary of a Report of 72 314 Cases From the Chinese Center for Disease Control and Prevention. JAMA 323, 1239 (2020).

6. Chinazzi, M. et al.. The effect of travel restrictions on the spread of the 2019 novel coronavirus (COVID-19) outbreak. Science eaba9757 (2020) doi:10.1126/science.aba9757.

7. Colbourn, T. COVID-19: extending or relaxing distancing control measures. The Lancet Public Health (2020) doi:10.1016/S2468-2667(20)30072-4.

8. Prem, K. et al.. The effect of control strategies to reduce social mixing on outcomes of the COVID-19 epidemic in Wuhan, China: a modelling study. The Lancet Public Health (2020) doi:10.1016/S2468-2667(20)30073-6.

9. Malta, M., Rimoin, A. W. & Strathdee, S. A. The coronavirus 2019-nCoV epidemic: Is hindsight 20/20? EClinicalMedicine 20, 100289 (2020).

10. Pan, A. et al.. Association of Public Health Interventions With the Epidemiology of the COVID-19 Outbreak in Wuhan, China. JAMA (2020) doi:10.1001/jama.2020.6130.

11. Mandal, S. et al.. Prudent public health intervention strategies to control the coronavirus disease 2019 transmission in India: A mathematical model-based approach. Indian Journal of Medical Research 0, 0 (2020).

12. Kucharski, A. J. et al.. Early dynamics of transmission and control of COVID-19: a mathematical modelling study. The Lancet Infectious Diseases (2020) doi:10.1016/S1473-3099(20)30144-4.

13. Li, Q. et al.. Early Transmission Dynamics in Wuhan, China, of Novel Coronavirus– Infected Pneumonia. New England Journal of Medicine 382, 1199–1207 (2020).

14. Kissler, S. M., Tedijanto, C., Goldstein, E., Grad, Y. H. & Lipsitch, M. Projecting the transmission dynamics of SARS-CoV-2 through the postpandemic period. Science (2020) doi:10.1126/science.abb5793.

15. Keeling, M. J. & Rohani, P. Modeling Infectious Diseases in Humans and Animals. (Princeton University Press, 2011).

16. Gani, R. & Leach, S. Transmission potential of smallpox in contemporary populations.Nature 414, 748–751 (2001).

17. Liu, Y., Gayle, A. A., Wilder-Smith, A. & Rocklöv, J. The reproductive number of COVID-19 is higher compared to SARS coronavirus. Journal of Travel Medicine 27, (2020).

18. Delamater, P. L., Street, E. J., Leslie, T. F., Yang, Y. T. & Jacobsen, K. H. Complexity of the Basic Reproduction Number (R ^0^). Emerging Infectious Diseases 25, 1–4 (2019).

19. Bauch, C. T., Lloyd-Smith, J. O., Coffee, M. P. & Galvani, A. P. Dynamically Modeling SARS and Other Newly Emerging Respiratory Illnesses: Past, Present, and Future. Epidemiology 16, 791–801 (2005).

20. Riley, S. Transmission Dynamics of the Etiological Agent of SARS in Hong Kong: Impact of Public Health Interventions. Science 300, 1961–1966 (2003).

21. Hellewell, J. et al.. Feasibility of controlling COVID-19 outbreaks by isolation of cases and contacts. The Lancet Global Health 8, e488–e496 (2020).

22. Viceconte, G. & Petrosillo, N. COVID-19 R0: Magic number or conundrum? Infectious Disease Reports 12, (2020).

23. Wearing, H. J., Rohani, P. & Keeling, M. J. Appropriate Models for the Management of Infectious Diseases. PLoS Medicine 2, e174 (2005).

24. Coronavirus. https://www.who.int/emergencies/diseases/novel-coronavirus-2019.

25. The COVID Tracking Project. The COVID Tracking Project https://covidtracking.com/about-data.

26. I.Stat Metadata Viewer. http://dati.istat.it/OECDStat_Metadata/ShowMetadata.ashx?Dataset=DCIS_POPRES1&ShowOnWeb=true&Lang=it.

27. CDC. Coronavirus Disease 2019 (COVID-19) in the U.S. Centers for Disease Control and Prevention https://www.cdc.gov/coronavirus/2019-ncov/cases-updates/cases-in-us.html (2020).

28. Provisional Death Counts for Coronavirus Disease (COVID-19). https://www.cdc.gov/nchs/nvss/vsrr/covid19/index.htm (2020).

29. MoHFW | Home. https://www.mohfw.gov.in/dashboard/index.php.

30. Lauer, S. A. et al.. The Incubation Period of Coronavirus Disease 2019 (COVID-19) From Publicly Reported Confirmed Cases: Estimation and Application. Annals of Internal Medicine (2020) doi:10.7326/M20-0504.

31. Patwardhan, S. C., Narasimhan, S., Jagadeesan, P., Gopaluni, B. & L. Shah, S. Nonlinear Bayesian state estimation: A review of recent developments. Control Engineering Practice 20, 933–953 (2012).

32. Kraemer, M. U. G. et al. The effect of human mobility and control measures on the COVID-19 epidemic in China. Science eabb4218 (2020) doi:10.1126/science.abb4218.

33. Newton, P. N. et al.. COVID-19 and risks to the supply and quality of tests, drugs, and vaccines. The Lancet Global Health (2020) doi:10.1016/S2214-109X(20)30136-4.

34. Buckee, C. O. et al.. Aggregated mobility data could help fight COVID-19. Science 368, 145.2–146 (2020).

35. The Lancet. The gendered dimensions of COVID-19. The Lancet 395, 1168 (2020).

36. Xu, B. et al.. Epidemiological data from the COVID-19 outbreak, real-time case information. Scientific Data 7, (2020).

37. Baud, D. et al.. Real estimates of mortality following COVID-19 infection. The Lancet Infectious Diseases (2020) doi:10.1016/S1473-3099(20)30195-X.

38. Wu, J. T. et al.. Estimating clinical severity of COVID-19 from the transmission dynamics in Wuhan, China. Nature Medicine (2020) doi:10.1038/s41591-020-0822-7.

